# Assessment of the COVID-19 Vaccine Program: Impact of the No Mask Mandate Executive Order in the State of Texas

**DOI:** 10.1101/2021.04.08.21255156

**Authors:** Ariana Richardson, Rachel Ruffin, Enahoro A. Iboi

## Abstract

On March 10, 2021, a new executive order to lift the mask mandate and allow businesses to open at 100 percent capacity, went into effect in the U.S. state of Texas. This was due to the decrease in the daily number of COVID-19 cases and deaths as the state continues to vaccinate the population. A simple compartmental model was used to assess the implications of the executive order on the ongoing vaccination program. Our simulation shows that approximately 51% of the entire population needs to be fully vaccinated to bring the control reproduction number to a value less than one (threshold condition needed for disease elimination) as compared to the 14.32% that has been fully vaccinated as of March 31, 2021. Hence, the need for an aggressive vaccination program if the state is to open businesses to full capacity and do not require the use of a face mask by the general public.

## 1 Introduction

The novel coronavirus (COVID-19) pandemic caused by SARS-CoV-2 continues to cause devastating public health and social-economic impact in the United States, including the state of Texas [1, 2]. This severe acute respiratory syndrome, is easily transmitted through person-to-person contact, most often through respiratory droplets when talking, coughing, or sneezing [3]. The United States, which is now the epicenter of the coronavirus outbreak, accounts for over 29,348,298 confirmed cases and 532,593 confirmed deaths (as of March 13, 2021) [4]. The US state of Texas as of March 13, 2021 accounts for over 2,715,993 confirmed cases, 46,007 confirmed deaths [4]. Non-pharmaceutical interventions (NPIs) to control the local transmission of the virus include contact tracing, quarantine, isolation, social-distancing, the use of face masks in public, and proper/regular washing of the hand [2, 5–9].

Pharmaceutical interventions such as a safe and effective vaccine is currently being administered aimed at containing COVID-19. The Pfizer, Inc., Moderna, Inc. and Janssen Biotech, Inc. third phase clinical trial that shows that their vaccine candidate which went through various stages of vaccine development have an estimated protective efficacy of about 95% (Pfizer, Inc.), 94.1% (Moderna Inc.), 66.3% overall, 74.4% effectiveness in the United States (Johnson & Johnson) and 76% (AstraZeneca) [10–13]. The Pfizer Inc. vaccine requires two doses separated by 21 days, and is administered by an intramuscular injection in the deltoid muscle [14]. The vaccine has been approved to those of 16 years of age and older. The Moderna Inc. vaccine is similar to the Pfizer vaccine, however these doses are separated by 28 days and administered to those of 18 years and older [15]. The J&J’s vaccine varies is a suspension for intramuscular injection administered as a single dose (0.5 mL). The dose is only ministered to those 18 years of age and older [13, 16].

Due to the limited supply of the vaccine to the general public, vaccine advisers to the US Centers for Disease Control and Prevention recommended that both health care workers and residents of long-term care facilities be first in line for any coronavirus vaccines [17]. On December 14, 2020, the US state of Texas was allocated 224,250 vaccine doses, marking the first week of vaccine distribution [18]. Texas has now become the first state to administer over one million vaccines, which is about half of the dosages they have received overall [19]. As of March 11, 2021, approximately 8.96% of the entire population are fully vaccinated in the state of Texas (see Figure 1). There has been decline in daily COVID-19 cases, hospitalization, and mortality which was attributed to the ongoing vaccination program in the state. Recently, the governor of the state of Texas lifted the mask mandate and allows businesses to open at 100% capacity due to the decline in COVID-19 cases, hospitalizations, and deaths [20].

**Figure 1:**
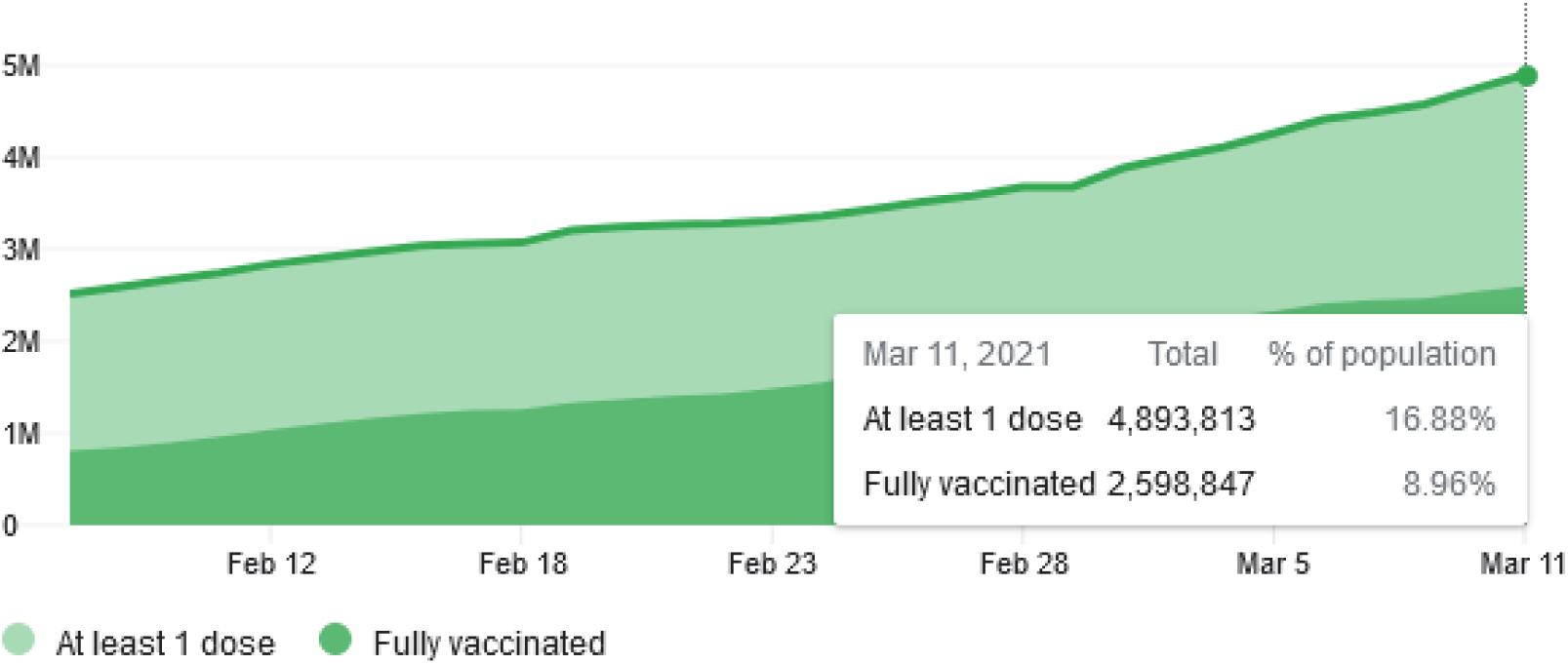
Data showing people who received at least 1 dose of a vaccine and people who are fully vaccinated in the US state of Texas as of March 11, 2021 [21].

Previous modeling studies have provided insights into public health measures for mitigating the spread of the novel coronavirus pandemic (see for instance [2, 5–8, 22–25]). The objective of this study is to use a mathematical model to assess the impact of the executive order to lift mask mandate and allow businesses to open at 100% capacity in the face of the ongoing coronavirus outbreak in the US state of Texas. The model is formulated in Section 2. The results are presented in Section 3 which include the asymptotic stability of the disease-free equilibrium of the model, parameters estimation of the model, and numerical simulations of the model. Discussions and concluding remarks are presented in Section 4.

## 2 Materials and Methods

### 2.1 Model Formulation

The model to be developed is for the transmission dynamics of the novel coronavirus pandemic in the US state of Texas. The total population at time *t*, denoted by *N* (*t*) is divided into into mutually exclusive compartments of unvaccinated susceptible individuals (*S*_*u*_(*t*)), vaccinated susceptible individuals (*S*_*v*_(*t*)), exposed individuals (*E*(*t*)), infectious with symptoms individuals (*I*_*s*_(*t*)), asymptomatic-infectious infectious individuals (*A*(*t*)), hospitalized or isolated (*H*(*t*)), individuals in intensive care units (*I*_*cu*_(*t*)), and recovered individuals (*R*(*t*)). Thus, the total population size *N* is given as

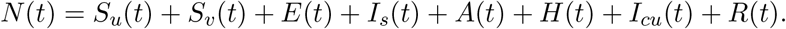

The model is given by the following deterministic system of nonlinear differential equations (where a dot represents differentiation with respect to time *t*):

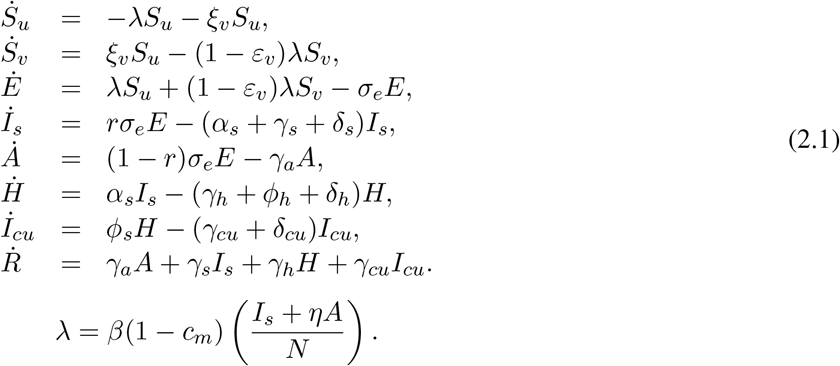

A flow diagram of the model is depicted in Figure 2. In model (2.1), the parameter *β* represent the effective contact rate and *c*_*m*_(0 *< c*_*m*_ ≤ 1) is the proportion of individuals who wear face masks (correctly and consistently) in public, while *η* is the modification parameters (0 *< η <* 1) that accounts for a reduction in infectiousness of individuals in *I*_*s*_ class. Since the vaccine is already being administered to front-line workers and people with pre-existing conditions, we assume that these people are vaccinated at a rate *ξ*_*v*_. We assume the vaccine induce protective efficacy 0 *< ε*_*v*_ < 1 in all individuals. Furthermore, the parameter *σ*_*e*_ represent the progression rate of exposed individuals. A proportion, 0 *< r ≤* 1, of exposed individuals show clinical symptoms of COVID-19 and move to the class *I*_*s*_ at the end of the incubation period. The remaining proportion, 1 *− r*, show no clinical symptoms and move to the *A* class. The parameter *α*_*s*_ is the hospitalization (or self-isolation) rate of individuals with clinical symptoms of COVID-19. Similarly, the parameter *ϕ*_*h*_ is the ICU admission rate. The parameters *γ*_*a*_, *γ*_*s*_, *γ*_*h*_, *γ*_*cu*_ represents the recovery rates for individuals in the *A, I*_*s*_, *I*_*cu*_ classes. Finally, the parameter *δ*_*s*_, *δ*_*h*_, *δ*_*cu*_ represents the COVID-induced mortality rate for individuals in the *I*_*s*_, *H, I*_*cu*_ classes. The model (2.1) is similar to other COVID-19 models in [2, 7, 8, 22, 23].

**Figure 2:**
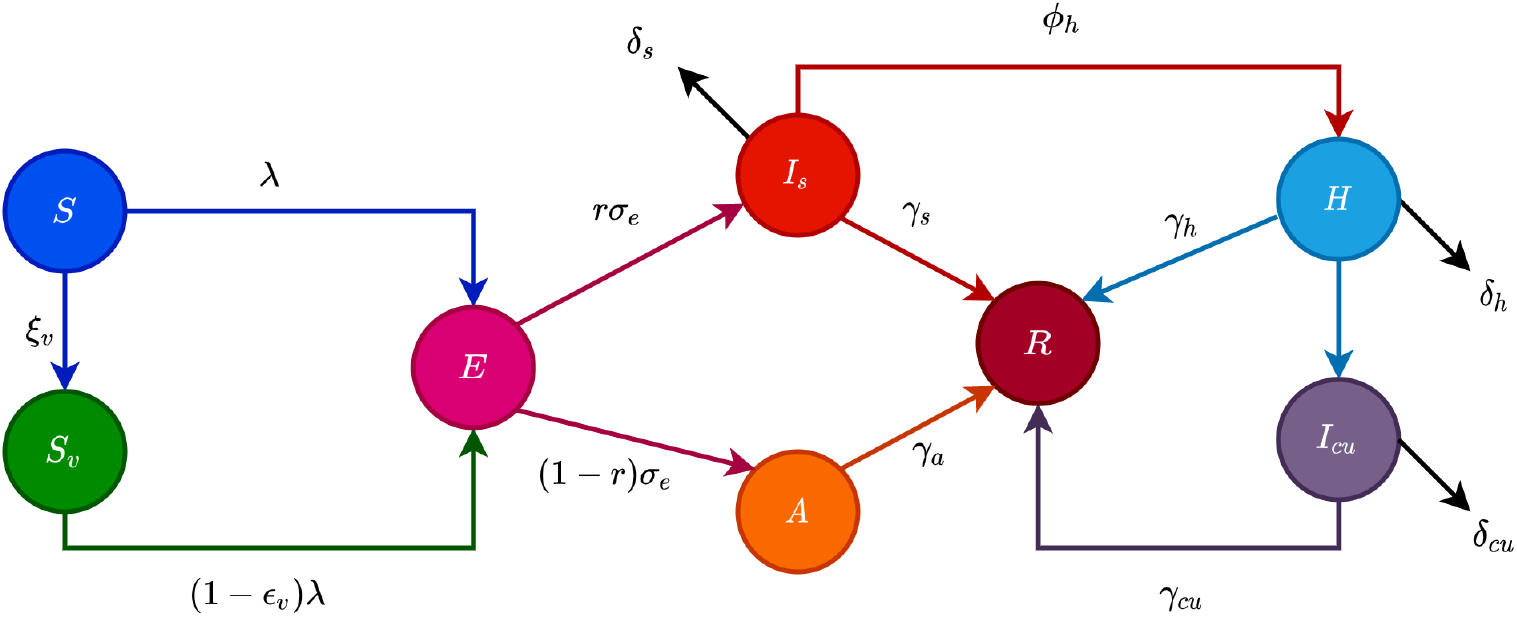
Flow diagram of the model (2.1) showing transitions from various compartments.

## 3 Results

### 3.1 Asymptotic Stability of Disease-free Equilibria

The closed form of the control reproduction number and the basic reproduction number in the absence of public health interventions are given in Appendix A. Using the parameter values in Table 2, for the period of January 12, 2021 to March 9, 2021, the control reproduction number for the state of Texas is 1.07 while the basic reproduction number is 1.56.

**Table 1:** Description of the state variables of the model (2.1)

| State variable | Description |
| --- | --- |
| $S_u$ | Population of unvaccinated susceptible individuals |
| $S_v$ | Population of vaccinated susceptible individuals |
| $E$ | Population of exposed individuals |
| $A$ | Population of asymptomatic-Infectious individuals |
| $I_s$ | Population of infectious individuals with severe clinical symptoms |
| $H$ | Population of hospitalized individuals |
| $I_{cu}$ | Population of individuals in ICU |
| $R$ | Population of recovered individuals |
| Parameter | Description |
| $\beta$ | Effective contact rates |
| $\xi_v$ | Efficacy of the vaccine in preventing COVID-19 infection ($0 < \xi_v \leq 1$) |
| $c_m$ | Face mask coverage ($0 < c_m \leq 1$) |
| $\eta$ | Modification parameters ($0 < \eta < 1$) |
| $\sigma_e$ | Progression rates from $E$ to $A$ or $I_s$ class |
| $r$ | Proportion of individuals in $E$ class who show clinical symptoms |
| $\alpha_s$ | Hospitalization rates for infectious individuals |
| $\phi_h$ | ICU admission rate for hospitalized individuals |
| $\gamma_a(\gamma_s)(\gamma_h)(\gamma_{cu})$ | Recovery rates for individuals |
| $\delta_s(\delta_h)(\delta_{cu})$ | Disease-induced death rates |

**Table 2:** Parameter values for the model (2.1).

| Parameter | Value | Reference |
| --- | --- | --- |
| $c_m$ | 0.31 | Assumed |
| $\sigma_e$ | 1/5.1 | [2, 22, 33, 34] |
| $r$ | 0.35 | [35, 36] |
| $\xi_v$ | 0.00299 | Estimated |
| $\varepsilon_v$ | 0.95 | [10, 11] |
| $\gamma_s$ | 1/10 | [5, 34] |
| $\gamma_a$ | 1/5 | [37] |
| $\gamma_h$ | 1/8 | [5] |
| $\gamma_{cu}$ | 1/10 | [5, 34] |
| $\eta$ | 0.25 | Assumed |
| $\beta$ | 0.2756 | Fitted |
| $\phi_h$ | 0.083 | Estimated |
| $\alpha_s$ | 0.025 | Estimated |
| $\delta_s$ | 0.02052 | Fitted |
| $\delta_h$ | 0.015 | Estimated |
| $\delta_{cu}$ | 0.025 | Estimated |

### 3.2 Data Fitting and Parameter Estimation

The model (2.1) uses estimates for some of the non vaccine related parameters obtained from the literature (as indicated in Table 2). Fitting the model (2.1) provided other parameter values in the context of a COVID-19 vaccine and the observed cumulative mortality data for the U.S. state of Texas. Specifically, we obtained from the John Hopkins University COVID-19 database, the cumulative mortality data from January 12, 2020 (first week of COVID-19 vaccination in Texas) to March 9, 2021 (1 day before the end of Texas’s mask mandate) [26]. We fit the model for this time period of the pandemic in order to correctly capture the trends observed in the cumulative mortality data leading up to the end of Texas’s mask mandate. Our decision to fit the model to mortality data, rather than case data, is due to the evidence of under-reporting and under-testing of COVID-19 cases in countries such as France, Italy, United States, Iran, and Spain [27] [7]. Mortality data therefore may provide a better indicator for COVID-19 case spread. The process of data-fitting involves applying the standard nonlinear least squares approach using the *fmincon* Optimization Toolbox embedded in MATLAB. The estimated values of the unknown parameters are tabulated in Table 2. Figure 3 depicts the fitting of the observed and predicted cumulative mortality for the U.S. state of Texas. Moreover, Figure 5 compares the simulations of the model using the fitted and fixed parameters in Table 2, to cumulative mortality, as a function of time, for various percentages of people who are fully vaccinated in Texas. The fitted model and these simulated data were used for the following identifiability analyses.

**Figure 3:**
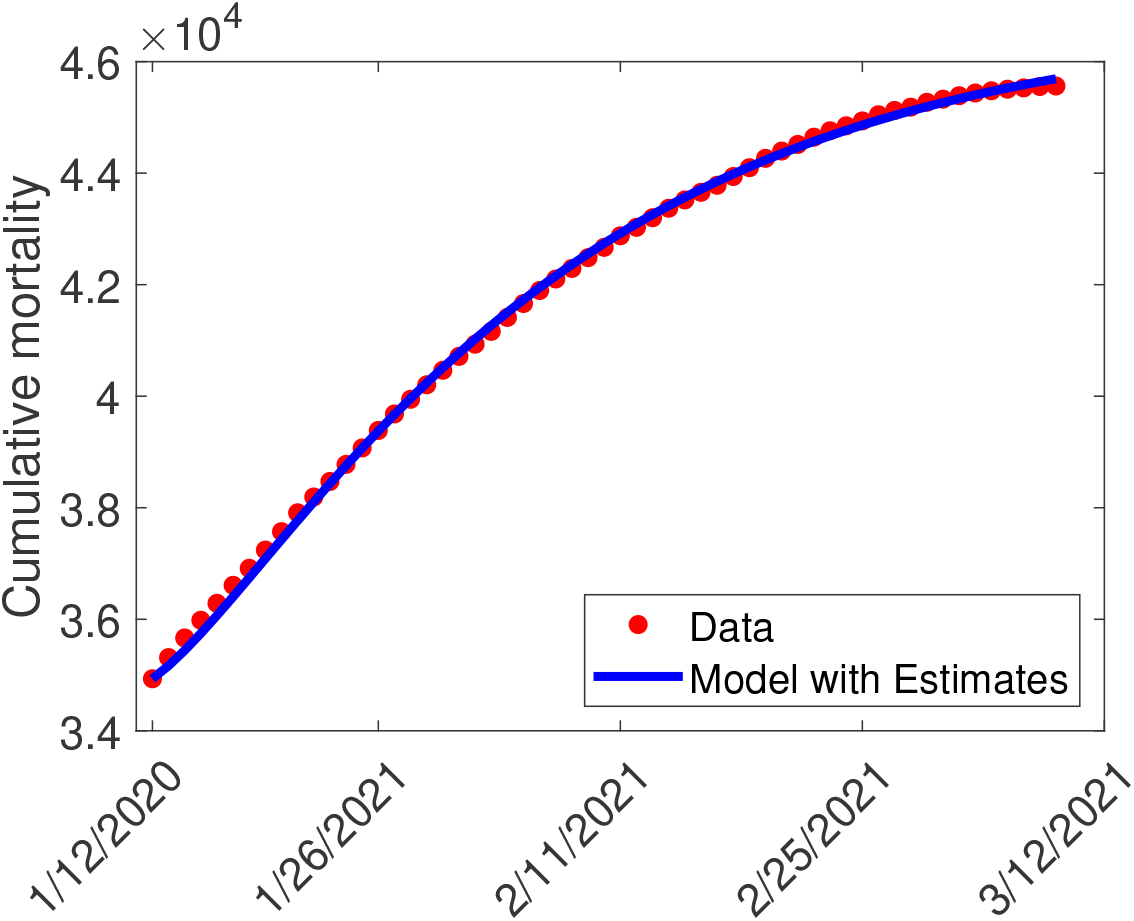
Data fitting of the model (2.1) using the cumulative mortality data for the US state of Texas from January 12, 2020 to March 9, 2021.

In order to address practical identifiability, we follow the profile likelihood approach in [28, 29]. The goal is to find confidence intervals for the estimated parameters *β* and *δ*_*s*_ respectively. Practical identifiability answers the question of whether unique parameter values can be estimated given available noisy data [30]. A confidence interval for the parameter of interest can be determined using the profile likelihood method. This is accomplished with the use of a threshold. The intersection points between the threshold and the profile likelihood curve at given parameter values result to the bounds of the 95% confidence interval [31]. A parameter is said to be practically identifiable if the shape of the profile likelihood plot is locally convex with a finite confidence interval as shown in Figure 4 [32].

**Figure 4:**
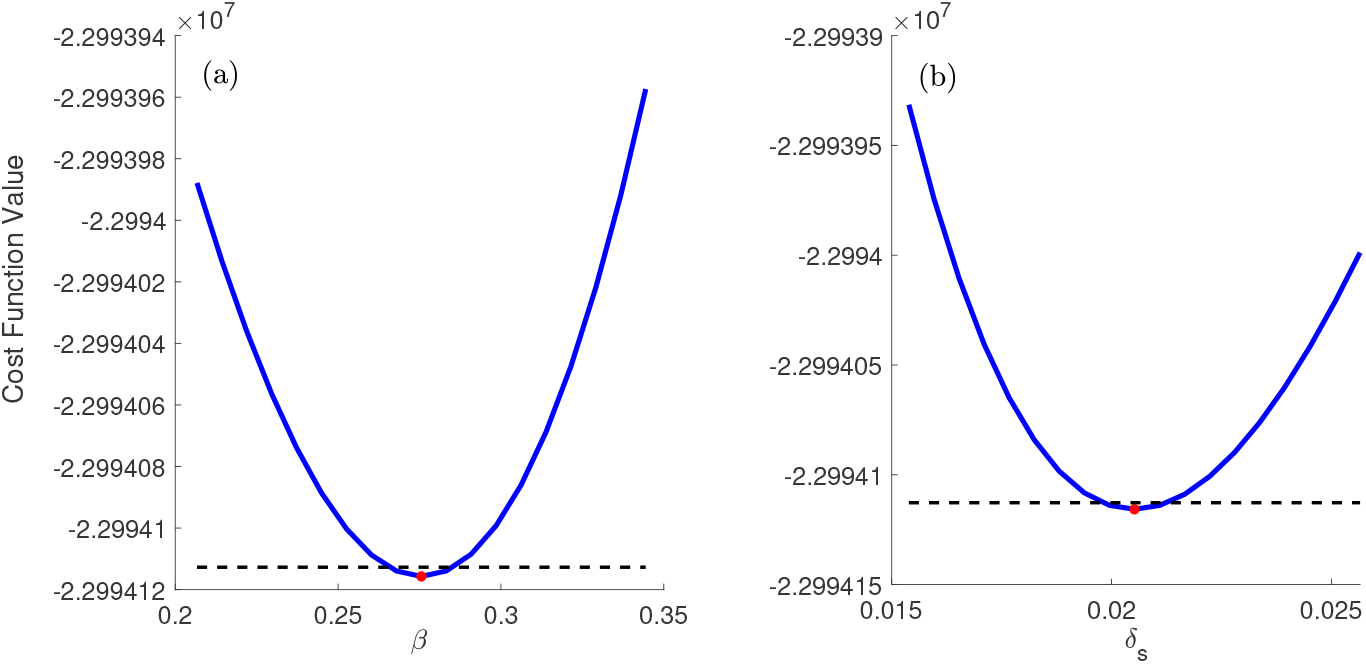
Profile likelihood plots for parameters *β* and *δ*_*s*_, while fixing the remaining parameters. The black dotted line depicts the threshold and the intersection points between the threshold and profile likelihood curve are the 95% confidence interval bounds with two degrees of freedom (d.f= 2). The red points are the parameter estimates shown in Table 2.

### 3.3 Numerical Simulation Results

The impact of the executive order (i.e., no mask mandate and businesses opening 100%) that went into effect on March 10, 2021, on the cumulative mortality in the state of Texas will now be explored through simulation of model 2.1 using parameter values in Table 2 unless otherwise stated. It is worth mentioning that since the Pfizer-BioNTech and Moderna vaccines are approximately 95% effective, then fully vaccinated individuals corresponds to having received both doses of the vaccine. Figure 5 shows the impact of increase in vaccination rate from the baseline value of 45,560 on the cumulative mortality. Our simulation result shows a projected 44,210 cumulative mortality with a 5-fold increase in vaccination rate (which corresponds to fully vaccinating 574,000 people daily) from the baseline value by March 9, 2021. This is a 3% decrease in cumulative mortality when compared to the baseline scenario. A similar trend was observed with a 10-fold increase in vaccination rate (which corresponds to fully vaccinating 1.2 million people daily) from the baseline value shows a projected 43,220 cumulative mortality by March 9, 2021. This is a 5.4% decrease in cumulative mortality when compared to the baseline scenario. Further, a 15-fold increase in vaccination rate (which corresponds to fully vaccinating 1.7 million people daily) from the baseline value shows a projected 42,630 cumulative mortality by March 9, 2021 which is a 7% decrease in the cumulative mortality from the baseline scenario. The result from Figure 5 supports the need for an aggressive vaccination program geared towards a drastic reduction in the daily COVID-19 induced mortality.

**Figure 5:**
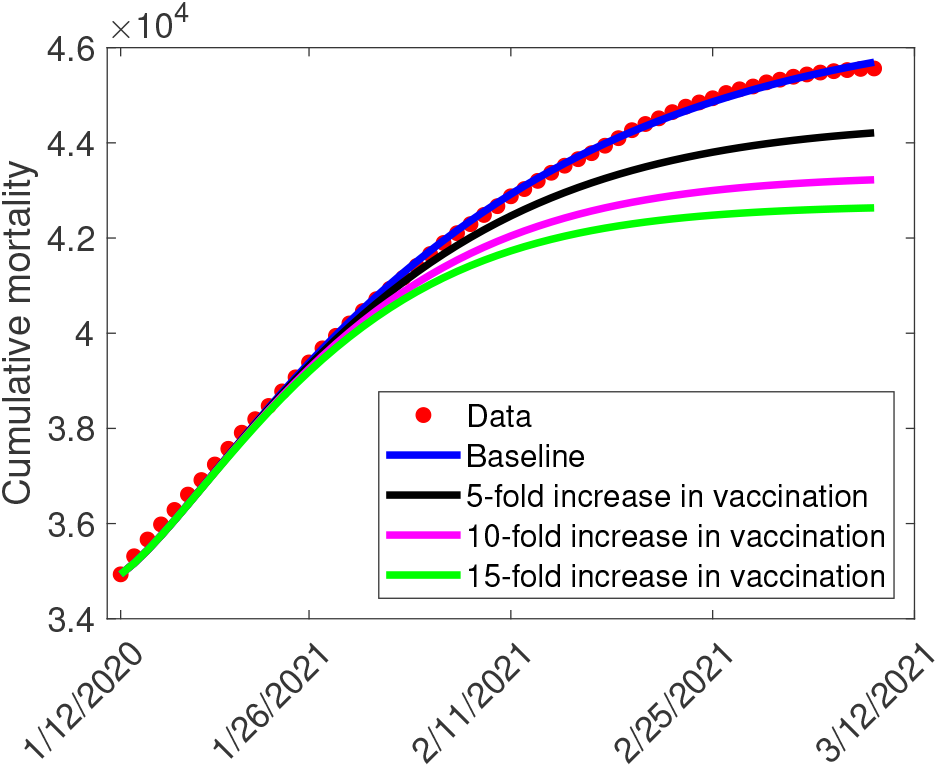
Simulations of the model, showing cumulative mortality, as a function of time, for various increase in the vaccination rates in the US state of Texas as at March 9, 2021. Solid dashed vertical lines depict the beginning of the no mask order. Other parameter values used for the simulations are as given in Tables 2.

A contour plot that shows the control reproduction number (ℛ _*c*_), as a function of the percentage of the population who are fully vaccinated and face mask coverage is depicted in Figure 6. Figure 6(a) for the baseline community contact rate shows that in the absence of face mask use, approximately 38% of the entire population needs to be fully vaccinated in order to bring the control reproduction number to a value less than one (threshold condition needed for disease elimination). However, as of March 28, 2021, approximately 13% of the entire population are fully vaccinated. The public health implication is that approximately 38% mask coverage is needed to complement the vaccination program in order to effectively control the spread of the virus (i.e., bring control reproduction number to a value less than one). Similarly, Figure 6(b) shows what could happen if businesses are to reopen 100% (this we assume corresponds to a 20% increase in the baseline value of the community contact rate parameter) and in the absence of face mask use. Our result shows that approximately 51% of the entire population needs to be fully vaccinated in order to bring the control reproduction number to a value less than one (threshold condition needed for disease elimination). This result further support the need for an aggressive vaccination program if the state is to open businesses 100% and do not require the use of a face mask by the general public.

**Figure 6:**
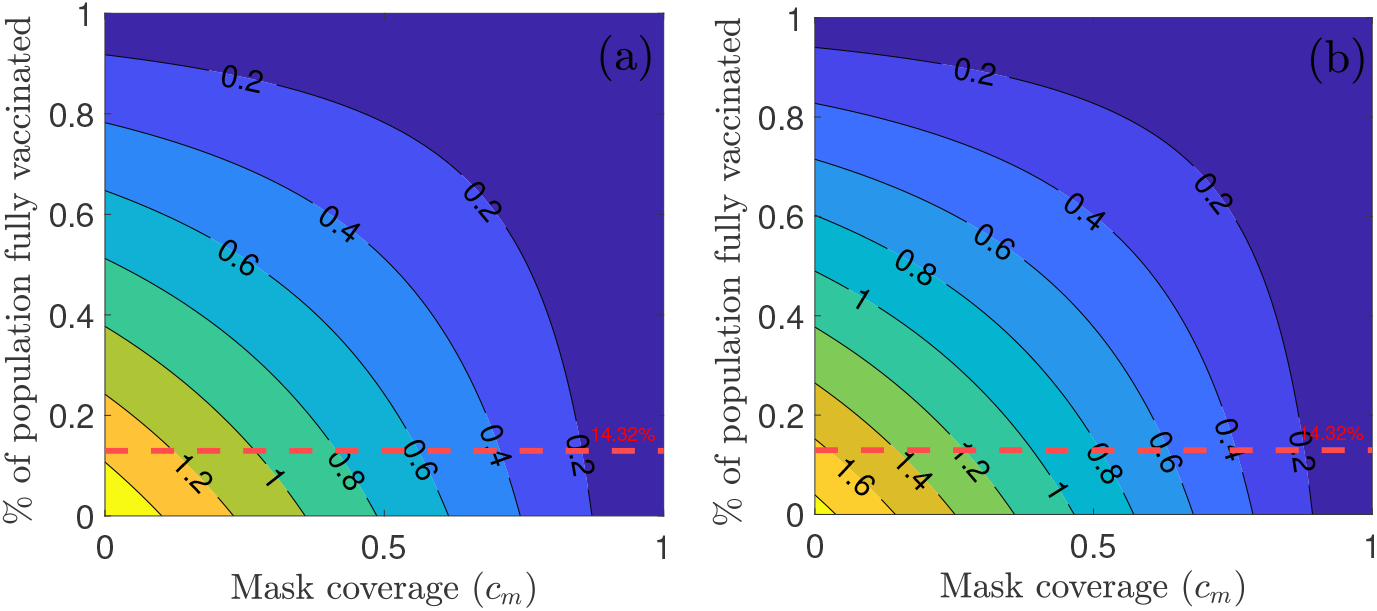
Contour plot of the control reproduction number (ℛ _*c*_) of the model, as a function of the % of population fully vaccinated and mask coverage (*c*_*m*_). (a) Baseline community contact rate parameter (*β*) (b) reopening of business 100% corresponds to a 20% increase in the baseline value of the community contact rate parameter (*β*). Red dashed horizontal lines depict the current 14.32% of population of Texas that are fully vaccinated as of March 31, 2021 [21]. Other parameter values used for the simulations are as given in Tables 2.

## 4 Summary and Conclusions

In order to understand the public health impact of the executive order issued in the state of Texas to allow businesses open to 100% capacity and to not require face mask use by the general public beginning from March 10, 2021 on the current vaccination program, we developed a COVID-19 model that captures the trends observed using the cumulative mortality data. The simulation shows the need for an aggressive vaccination program to reduce COVID-19 induced mortality. For example, a 15-fold increase in vaccination rate (which corresponds to fully vaccinating 1.7 million people daily) from the baseline value shows a projected 42,630 cumulative mortality by March 9, 2021 which is a 7% decrease in the cumulative mortality from the baseline scenario. We assessed the impact of combining the routine vaccination strategy with a public mask use strategy. Our result shows that 38% mask coverage is needed to complement the vaccination program in order to effectively control the spread of the virus in the community. However, if businesses are to reopen 100% (this we assume corresponds to a 20% increase in the baseline value of the community contact rate parameter) and face mask is no longer required in public places, our result shows that approximately 51% of the entire population needs to be fully vaccinated in order to bring the control reproduction number to a value less than one. This is consistent with the result obtained in Iboi *et al*. [8]. In summary, this study shows the importance of complementing the recent success in the fight to eliminating COVID-19 in the state of Texas through vaccination with other non-pharmaceutical interventions such as the use of face mask and practising social distance. The relaxation of these interventions should to be a gradual process and not a one time order so as not to jeopardize the ongoing reduction in daily cases and deaths being recorded.

## Data Availability

https://github.com/CSSEGISandData/COVID-19

https://github.com/owid/covid-19-data/blob/master/public/data/vaccinations/us_state_vaccinations.csv

## Acknowledgements

AR, RR, and EAI would like to acknowledge and thank our partners of The Boeing Company and the Thurgood Marshall College Fund, for their charitable, capacity grant in support of the Math RaMP Program at Spelman College, via the Boeing | TMCF HBCU Investment. We would also like to acknowledge the MAA Tensor Women and Mathematics Program for support of the Math RaMP Program at Spelman College. EAI would like to acknowledge the support of the National Science Foundation (Award #1761945).

## Authors contributions

EAI, AR, and RR conceived the study; EAI, AR, and RR designed the model; AR and RR collected and analysed the data; EAI performed the numerical simulations; AR, and RR drafted the manuscript. All authors read and approved the final manuscript. Ethics approval was not required as all data used in the manuscript are publicly available.

## Conflicts of Interest

All authors declare no conflicts of interest.

## A Tables of variable descriptions, parameter descriptions, and parameter values

## Appendix A Control Reproduction Number for the Model

The model (2.1) has a continuum (or line) of disease-free equilibria (DFE), given by

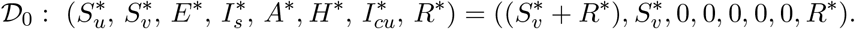

where *N*^∗^ is the initial total population size, 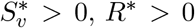, and 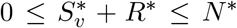. The next generation operator method [38, 39] can be used to analyse the asymptotic stability property of the continuum of disease-free equilibria, 𝒟_0_. In particular, using the notation in [38], it follows that the associated next generation matrices, *F* and *V*, for the new infection terms and the transition terms, are given, respectively, by

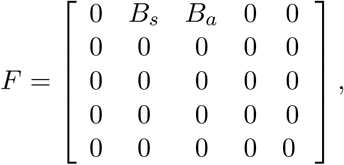

and,

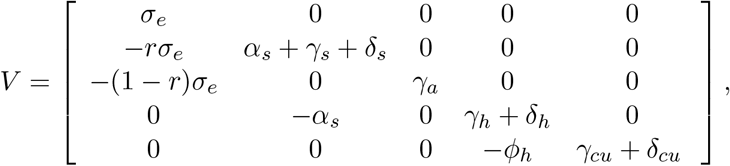

where,

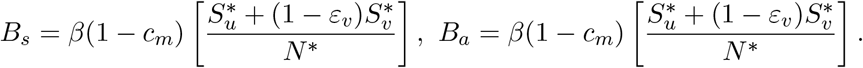

It is convenient to define the quantity *ℛ*_*c*_ by:

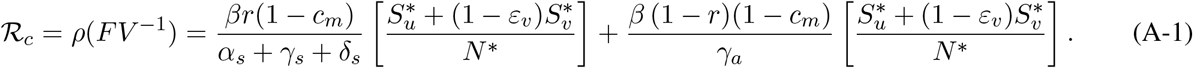

where *ρ* is the spectral radius.

The quantity ℛ_*c*_ is the *control reproduction number* of the model (2.1). It measures the average number of new COVID-19 cases generated by a typical infectious individual introduced into a population where a certain fraction is protected. It is the sum of the constituent reproduction numbers associated with the number of new cases generated by symptomatically-infectious humans 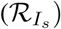 and asymptomatically-infectious (ℛ_*A*_) individuals. In the absence of public health measure, the basic reproduction number of the model (2.1) is given by

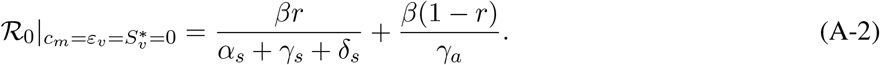

## Notes

### Competing Interest Statement

The authors have declared no competing interest.

### Clinical Trial

n/a

## References

[1] W. H. Organization, “Coronavirus Disease (COVID-19) Dashboard,” (2020 (accessed October 30, 2020)).

[2] C. N. Ngonghala, E. Iboi, S. Eikenberry, M. Scotch, C. R. MacIntyre, M. H. Bonds, and A. B. Gumel, “Mathematical assessment of the impact of non-pharmaceutical interventions on curtailing the 2019 novel coronavirus,” Mathematical Biosciences 108364 (2020).

[3] “Symptoms of coronavirus,” (2020).

[4] John Hopkins University, “Daily state-by-state testing trends,” John Hopkins University Coronavirus resource center (Assessed on October 30, 2020).

[5] N. M. Ferguson, D. Laydon, G. Nedjati-Gilani, N. Imai, K. Ainslie, M. Baguelin, S. Bhatia, A. Boonyasiri, Z. Cucunubá, G. Cuomo-Dannenburg, et al., “Impact of non-pharmaceutical interventions (NPIs) to reduce COVID-19 mortality and healthcare demand,” London: Imperial College COVID-19 Response Team, March 16 (2020).

[6] S. E. Eikenberry, M. Mancuso, E. Iboi, T. Phan, K. Eikenberry, Y. Kuang, E. Kostelich, and A. B. Gumel, “To mask or not to mask: Modeling the potential for face mask use by the general public to curtail the covid-19 pandemic,” arXiv preprint 2004.03251 (2020).

[7] C. N. Ngonghala, E. A. Iboi, and A. B. Gumel, “Could masks curtail the post-lockdown resurgence of covid-19 in the US?” Mathematical biosciences 329, 108452 (2020).

[8] E. A. Iboi, C. N. Ngonghala, and A. B. Gumel, “Will an imperfect vaccine curtail the covid-19 pandemic in the US?” Infectious Disease Modeling 5, 510–524 (2020).

[9] E. Iboi, A. Richardson, R. Ruffin, D. Ingram, J. Clark, J. Hawkins, M. McKinney, N. Horne, R. Ponder, Z. Denton, et al., “Impact of public health education program on the novel coronavirus outbreak in the United States,” Frontiers in Public Health 9, 208 (2021).

[10] “Pfizer and Biontech to submit emergency use authorization request today to the U.S. FDA for COVID-19 vaccine,” (2020).

[11] “Promising interim results from clinical trial of nih-moderna COVID-19 vaccine,” (2020).

[12] “Azd1222 vaccine met primary efficacy endpoint in preventing COVID-19,” (2020).

[13] “Janssen COVID-19 Vaccine,” (2021).

[14] “Pfizer-Biontech COVID-19 Vaccine,” (2021).

[15] “Moderna COVID-19 Vaccine,” (2021).

[16] “Janssen COVID-19 Vaccine (Johnson Johnson),” (2021).

[17] “Health care workers and long-term care facility residents should get covid-19 vaccine first, cdc vaccine advisers say,” (2020).

[18] “COVID-19 Vaccine Arrives in Texas,” (2021).

[19] “Texas becomes first state to administer 1 million doses of COVID-19 vaccine,” (2021).

[20] “Texas governor lifts mask mandate and allows businesses to open at 100% capacity, despite health officials’ warnings,” (2021).

[21] “Vaccinations,” (2021).

[22] S. E. Eikenberry, M. Muncuso, E. Iboi, T. Phan, E. Kostelich, Y. Kuang, and A. B. Gumel, “To mask or not to mask: Modeling the potential for face mask use by the general public to curtail the covid-19 pandemic,” Infectious Disease Modeling 5, 293–308 (2020).

[23] E. A. Iboi, O. O. Sharomi, C. N. Ngonghala, and A. B. Gumel, “Mathematical modeling and analysis of covid-19 pandemic in Nigeria,” Mathematical Biosciences and Engineering 17, 7192–7220 (2020).

[24] K. Mizumoto, K. Kagaya, A. Zarebski, and G. Chowell, “Estimating the asymptomatic proportion of coronavirus disease 2019 (COVID-19 cases on board the Diamond Princess cruise ship, Yokohama, Japan, 2020,” Eurosurveillance 25, 2000180 (2020).

[25] K. Mizumoto and G. Chowell, “Transmission potential of the novel coronavirus (COVID-19) onboard the Diamond Princess Cruises Ship, 2020,” Infectious Disease Modelling (2020).

[26] “Center for Systems Science and Engineering at Johns Hopkins University. COVID-19,” (2020).

[27] H. Lau, T. Khosrawipour, P. Kocbach, H. Ichii, J. Bania, and V. Khosrawipour, “Evaluating the massive underreporting and undertesting of COVID-19 cases in multiple global epicenters,” Pulmonology (2020).

[28] S. A. Murphy and A. W. V. D. Vaart, “On profile likelihood,” Journal of the American Statistical Association 95, 449–465 (2000).

[29] D. J. Venzon and S. H. Moolgavkar, “A method for computing profile-likelihood-based confidence intervals,” Journal of the Royal Statistical Society. Series C (Applied Statistics) 37, 87–94 (1988).

[30] J. A. Jacquez and P. Greif, “Numerical parameter identifiability and estimability: Integrating identifiability, estimability, and optimal sampling design,” Mathematical Biosciences 77, 201–227 (1985).

[31] M. T. B. J. S. M. K. U. T. J. Raue A, Kreutz C, “Structural and practical identifiability analysis of partially observed dynamical models by exploiting the profile likelihood,” Bioinformatics 25, 1923–9 (2009).

[32] J. H. V. Eisenberg, M. C., “A confidence building exercise in data and identifiability: Modeling cancer chemotherapy as a case study,” Journal of theoretical biology 431, 63–78 (2017).

[33] C. Zhou, “Evaluating new evidence in the early dynamics of the novel coronavirus COVID-19 outbreak in Wuhan, China with real time domestic traffic and potential asymptomatic transmissions,” medRxiv (2020).

[34] L. Zou, F. Ruan, M. Huang, L. Liang, H. Huang, Z. Hong, J. Yu, M. Kang, Y. Song, J. Xia, et al., “SARS-CoV-2 viral load in upper respiratory specimens of infected patients,” New England Journal of Medicine 382, 1177–1179 (2020).

[35] World Health Organization, “Coronavirus disease 2019 (COVID-19): situation report, 46,” WHO (2020).

[36] Z. Wu and J. M. McGoogan, “Characteristics of and important lessons from the coronavirus disease 2019 (COVID-19) outbreak in China: summary of a report of 72 314 cases from the Chinese Center for Disease Control and Prevention,” JAMA (2020).

[37] S. Kissler, C. Tedijanto, E. Goldstein, Y. Grad, and M. Lipsitch, “Projecting the transmission dynamics of SARS-CoV-2 through the postpandemic period,” Science (2020).

[38] P. van den Driessche and J. Watmough, “Reproduction numbers and sub-threshold endemic equilibria for compartmental models of disease transmission,” Mathematical Biosciences 180, 29–48 (2002).

[39] O. Diekmann, J. A. P. Heesterbeek, and J. A. Metz, “On the definition and the computation of the basic reproduction ratio R0 in models for infectious diseases in heterogeneous populations,” Journal of Mathematical Biology 28, 365–382 (1990).

